# Persistent Sex Disparities in Pre-hospital Delay Among STEMI Patients Despite Overall Improvements: Findings from the Chinese Cardiovascular Association Chest Pain Center Registry

**DOI:** 10.1101/2025.07.07.25331066

**Authors:** Zhenzhen Lu, Tongtong Zang, Hao Li, Yiling Liu, Li Shen, Lihong Huang

## Abstract

**BACKGROUND:** Women with ST-elevation myocardial infarction (STEMI) historically experience longer pre-hospital delays than men. Whether this disparity has narrowed in China following nationwide quality initiatives remains unknown.

**METHODS:** We analyzed 1,104,812 STEMI patients from the Chinese Cardiovascular Association Database-Chest Pain Center (2016–2023). Pre-hospital delay (symptom-to-door time) and its components (symptom-to-call [EMS call delay], call-to-door [transport delay]) were analyzed. Sex differences were assessed using multivariable quantile regression (adjusting for age, comorbidities, cardiovascular risk factors), examining trends over time (2016–2019, 2020–2022, 2023).

**RESULTS:** Women consistently experienced longer pre-hospital delays than men (median: 170 vs. 124 minutes; adjusted median difference: 24.44 min, 95%CI:22.71– 26.16, *P*<0.001). Although overall pre-hospital delay decreased from 2016 to 2023, the sex disparities persisted, particularly at higher delay percentiles (e.g., >60 min wider at 90th percentile by 2023). Decomposition revealed the disparity was primarily driven by EMS call delay (adjusted difference: 13.42 min, 95%CI:10.76–16.08, *P*<0.001), not transport delay (1.49 min, *P*<0.001). Younger women (<60 years) exhibited the largest relative disparity. The 2020–2022 pandemic period disrupted improvement trends, especially for EMS call delay.

**CONCLUSIONS:** Despite overall reductions in pre-hospital delay for STEMI in China, sex disparities persisted, especially younger women. Targeted interventions addressing symptom recognition and help-seeking behavior in women are urgently needed.

---

ST-elevation myocardial infarction (STEMI) necessitates rapid revascularization to minimize myocardial damage and mortality. Pre-hospital delay, defined as the time from symptom onset to hospital arrival, critically impacts outcomes, with each minute of delay increasing infarct size and worsening prognosis.^1,2^ Numerous studies have highlighted significant sex disparities in pre-hospital delay, with women often experiencing longer delays compared to men.^3-6^ These disparities are evident not only in the total pre-hospital delay but also in specific components such as emergency medical services (EMS) call delay (the time from symptom onset to calling emergency medical services) and EMS call-to-hospital arrival time (transportation delay).^5^ These delays are attributed to factors including atypical symptom presentation, socioeconomic barriers, cultural factors, and differences in healthcare-seeking behaviors. Women tend to present with STEMI at older ages and with more comorbidities, which can exacerbate delays in recognition and treatment.^5-9^_2,10_

While some developed healthcare systems report narrowing sex disparities in STEMI care timelines following systemic improvements,^11^ evidence from large, developing nations like China remains scarce. China has implemented a major national quality initiative — the Chest Pain Center accreditation — since 2016, aiming to standardize and accelerate STEMI care pathways.^12^ Whether this large-scale system improvement has translated into a reduction, or potentially elimination, of the sex disparity in pre-hospital delay is unknown. Understanding the current state and temporal trends of this disparity is crucial for designing equitable interventions.

Using data from the national Chinese Cardiovascular Association (CCA) Database — Chest Pain Center, this study aimed to: 1) Evaluate sex differences in pre-hospital delay and its components (EMS call delay, transport delay) among STEMI patients in China, and 2) Assess how these differences have evolved from 2016 to 2023.

## METHODS

The data used in this study were sourced from the CCA Database — Chest Pain Center. Detailed information on this database has been published previously.^13^ Data quality is monitored by the registry’s data management committee, and participating centers undergo accreditation to ensure standardized STEMI care protocols.

All data utilized in this study have undergone anonymization procedures and adhere to a privacy and confidentiality agreement. The anonymized data will be accessed upon formal request and subsequent review by the data management committee of the CCA Database — Chest Pain Center. The study was approved by the Ethical Committee of Zhongshan hospital, Fudan University (approval no.B2024-045).

### Study Design and Participants

This is an observational cohort study of STEMI patients recorded in the CCA Database — Chest Pain Center registry between January 1, 2016 and December 31, 2023. Eligible patients were adults (age ≥18) diagnosed with STEMI according to contemporaneous guidelines, typically requiring ischemic symptoms, diagnostic ST-segment elevations (or new left bundle branch block) on ECG, and elevated cardiac biomarkers. We excluded patients if their symptom onset occurred after hospital admission (i.e., inpatients), if they were immediately transferred out to another facility, or if their recorded pre-hospital delay exceeded 24 hours (to focus on acute presentations).

For analyses of EMS call delay, we further restricted the cohort to patients who arrived at the hospital via EMS (ambulance). Patients who self-transported were excluded from the EMS call delay subgroup analysis, since their EMS activation times were not applicable or not recorded.

### Variables

Pre-hospital delay was defined as the time (in minutes) from symptom onset to hospital arrival (door time). EMS call delay was defined as the interval from symptom onset to contacting EMS (i.e. phone call for an ambulance), and transportation delay as the time from EMS contact to hospital arrival. These time intervals were recorded by the receiving hospital based on patient reports and EMS records.

Baseline variables included age, sex, cardiovascular risk factors (history of hypertension, diabetes, hyperlipidemia, smoking status), previous cardiovascular disease, prior heart failure, and Killip class on presentation. Hospital-level factors (tertiary vs non-tertiary center) and region (East/Central/West China, as detailed in Supplementary Table S1) were noted, as well as mode of transport (EMS or self-transport). To quantify overall comorbidity burden, we calculated the Charlson Comorbidity Index for each patient based on their medical history (see Supplementary Table S2 for index components).^14^

### Statistical Analysis

Categorical variables were summarized as counts with percentages and compared using the chi-square tests. Continuous variables were expressed as means with standard deviations (SD) if approximately normally distributed and compared using the t-test; otherwise, they were summarized as medians with interquartile ranges (IQR) and compared using univariable quantile regression.

Temporal trends were evaluated by grouping patients into three admission periods: 2016–2019 (pre-COVID baseline), 2020–2022 (COVID-19 pandemic period), and 2023 (post-pandemic). We chose quantile regression for the primary analyses because delay times were highly skewed and we were specifically interested in differences across the distribution (not just the mean), including at extreme percentiles. Multivariable quantile regression models were constructed to estimate adjusted sex differences in delays at the 10th, 25th, 50th (median), 75th, and 90th percentiles of the time distribution. Ordinary least squares (OLS) linear regression was also performed for comparison. All multivariable regression models adjusted for age, cardiovascular risk factors, and comorbidities (summarized as a comorbidity index) to control for potential confounding effects. To assess whether sex disparities changed over time, we included an interaction term between sex and the admission period in the models; a significant interaction would indicate a different trend in delay for women compared to men across the time periods.

Subgroup analyses explored variations in pre-hospital delay across age groups (18–59, 60–79, ≥80 years), hospital levels (tertiary vs. non-tertiary), hospital regions (East, Central, West China), and patient arrival modes (EMS transport vs. self-transport).

Missing data were imputed using multiple imputation by chained equations (MICE), with details provided in Supplementary Table S3.

All analyses were conducted using R software (version 4.4.3, R Foundation). Statistical significance was set at a two-sided *P* ≤ 0.05.

## RESULTS

### Study Population

A total of 1,104,812 patients with STEMI were included in the analysis, comprising 854,774 men (77.4%) and 250,038 women (22.6%). Figure 1 shows the study flow diagram, including all inclusions and exclusions, and delineates the subset of patients who arrived via EMS (which was used for the EMS call delay and transportation delay analyses). Baseline characteristics stratified by sex are presented in Supplementary Table S4. Overall, female patients were significantly older than male patients (mean age 69.8 vs 60.0 years) and had higher rates of hypertension (55.8% vs 45.1%), diabetes mellitus (28.0% vs 20.0%), and heart failure on presentation (Killip class III–IV in 12.3% vs 7.9%). In contrast, women were far less likely to report smoking habits than men (4.9% vs 43.3%). Table 1 summarizes baseline characteristics stratified by sex and time period. Notably, both women and men in the later cohorts (2020–2022 and 2023) had a higher burden of cardiovascular risk factors (hypertension, diabetes, and hyperlipidemia) compared to those in the 2016–2019. The proportion of patients presenting to non-tertiary hospitals also increased over time for both sexes.

**Table 1.**
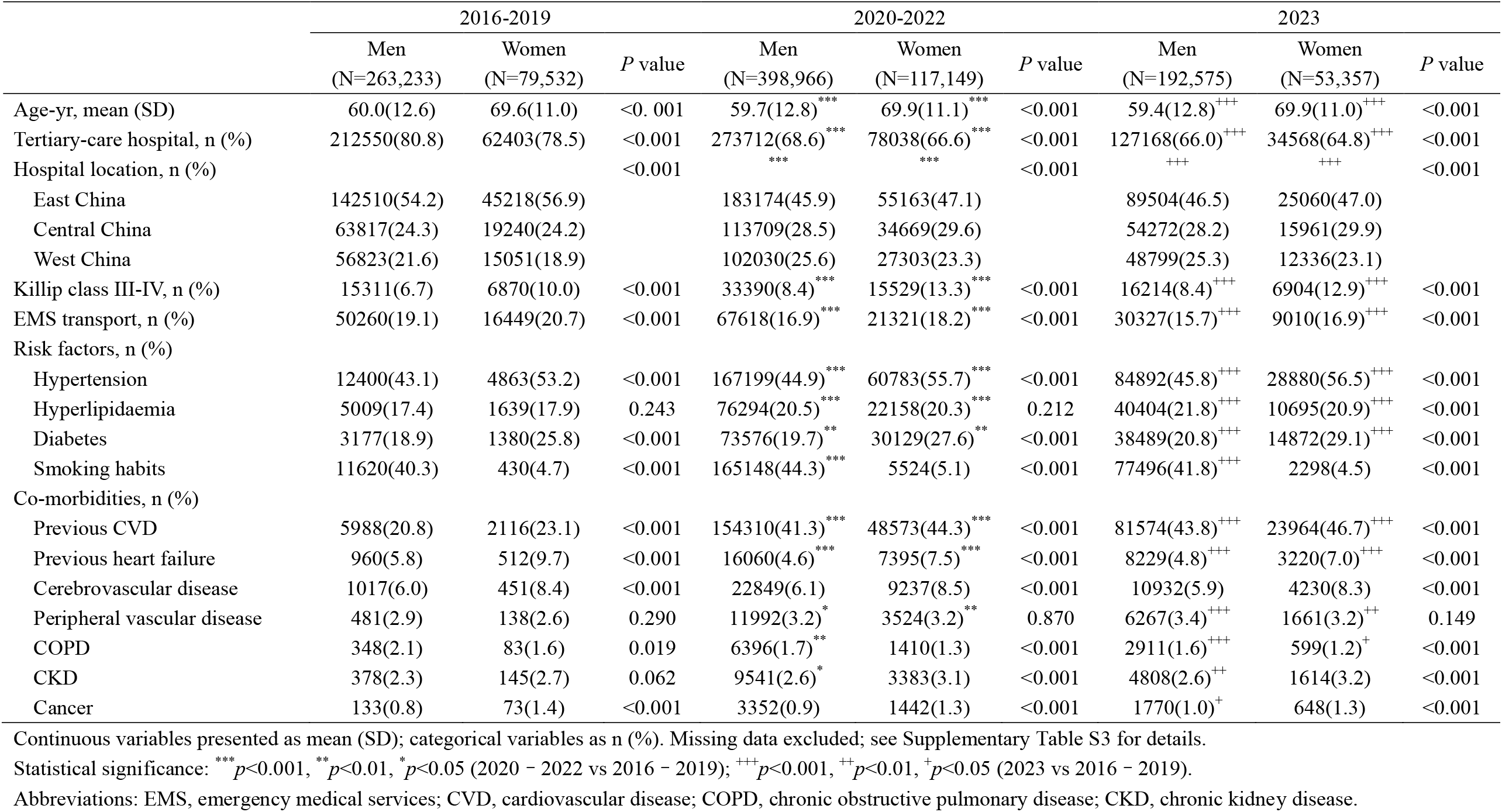
Baseline characteristics of STEMI patients, by sex and study period (2016–2023).

|  | 2016-2019 |  |  | 2020-2022 |  |  | 2023 |  |  |
| --- | --- | --- | --- | --- | --- | --- | --- | --- | --- |
|  | Men<br>(N=263,233) | Women<br>(N=79,532) | *P* value | Men<br>(N=398,966) | Women<br>(N=117,149) | *P* value | Men<br>(N=192,575) | Women<br>(N=53,357) | *P* value |
| Age-yr, mean (SD) | 60.0(12.6) | 69.6(11.0) | <0.001 | 59.7(12.8)<sup>***</sup> | 69.9(11.1)<sup>***</sup> | <0.001 | 59.4(12.8)<sup>+++</sup> | 69.9(11.0)<sup>+++</sup> | <0.001 |
| Tertiary-care hospital, n (%) | 212550(80.8) | 62403(78.5) | <0.001 | 273712(68.6)<sup>***</sup> | 78038(66.6)<sup>***</sup> | <0.001 | 127168(66.0)<sup>+++</sup> | 34568(64.8)<sup>+++</sup> | <0.001 |
| Hospital location, n (%) |  |  | <0.001 |  |  | <0.001 |  |  | <0.001 |
| East China | 142510(54.2) | 45218(56.9) |  | 183174(45.9) | 55163(47.1) |  | 89504(46.5) | 25060(47.0) |  |
| Central China | 63817(24.3) | 19240(24.2) |  | 113709(28.5) | 34669(29.6) |  | 54272(28.2) | 15961(29.9) |  |
| West China | 56823(21.6) | 15051(18.9) |  | 102030(25.6) | 27303(23.3) |  | 48799(25.3) | 12336(23.1) |  |
| Killip class III-IV, n (%) | 15311(6.7) | 6870(10.0) | <0.001 | 33390(8.4)<sup>***</sup> | 15529(13.3)<sup>***</sup> | <0.001 | 16214(8.4)<sup>+++</sup> | 6904(12.9)<sup>+++</sup> | <0.001 |
| EMS transport, n (%) | 50260(19.1) | 16449(20.7) | <0.001 | 67618(16.9)<sup>***</sup> | 21321(18.2)<sup>***</sup> | <0.001 | 30327(15.7)<sup>+++</sup> | 9010(16.9)<sup>+++</sup> | <0.001 |
| Risk factors, n (%) |  |  |  |  |  |  |  |  |  |
| Hypertension | 12400(43.1) | 4863(53.2) | <0.001 | 167199(44.9)<sup>***</sup> | 60783(55.7)<sup>***</sup> | <0.001 | 84892(45.8)<sup>+++</sup> | 28880(56.5)<sup>+++</sup> | <0.001 |
| Hyperlipidaemia | 5009(17.4) | 1639(17.9) | 0.243 | 76294(20.5)<sup>***</sup> | 22158(20.3)<sup>***</sup> | 0.212 | 40404(21.8)<sup>+++</sup> | 10695(20.9)<sup>+++</sup> | <0.001 |
| Diabetes | 3177(18.9) | 1380(25.8) | <0.001 | 73576(19.7)<sup>**</sup> | 30129(27.6)<sup>**</sup> | <0.001 | 38489(20.8)<sup>+++</sup> | 14872(29.1)<sup>+++</sup> | <0.001 |
| Smoking habits | 11620(40.3) | 430(4.7) | <0.001 | 165148(44.3)<sup>***</sup> | 5524(5.1) | <0.001 | 77496(41.8)<sup>+++</sup> | 2298(4.5) | <0.001 |
| Co-morbidities, n (%) |  |  |  |  |  |  |  |  |  |
| Previous CVD | 5988(20.8) | 2116(23.1) | <0.001 | 154310(41.3)<sup>***</sup> | 48573(44.3)<sup>***</sup> | <0.001 | 81574(43.8)<sup>+++</sup> | 23964(46.7)<sup>+++</sup> | <0.001 |
| Previous heart failure | 960(5.8) | 512(9.7) | <0.001 | 16060(4.6)<sup>***</sup> | 7395(7.5)<sup>***</sup> | <0.001 | 8229(4.8)<sup>+++</sup> | 3220(7.0)<sup>+++</sup> | <0.001 |
| Cerebrovascular disease | 1017(6.0) | 451(8.4) | <0.001 | 22849(6.1) | 9237(8.5) | <0.001 | 10932(5.9) | 4230(8.3) | <0.001 |
| Peripheral vascular disease | 481(2.9) | 138(2.6) | 0.290 | 11992(3.2)<sup>*</sup> | 3524(3.2)<sup>**</sup> | 0.870 | 6267(3.4)<sup>+++</sup> | 1661(3.2)<sup>++</sup> | 0.149 |
| COPD | 348(2.1) | 83(1.6) | 0.019 | 6396(1.7)<sup>**</sup> | 1410(1.3) | <0.001 | 2911(1.6)<sup>+++</sup> | 599(1.2)<sup>+</sup> | <0.001 |
| CKD | 378(2.3) | 145(2.7) | 0.062 | 9541(2.6)<sup>*</sup> | 3383(3.1) | <0.001 | 4808(2.6)<sup>++</sup> | 1614(3.2) | <0.001 |
| Cancer | 133(0.8) | 73(1.4) | <0.001 | 3352(0.9) | 1442(1.3) | <0.001 | 1770(1.0)<sup>+</sup> | 648(1.3) | <0.001 |

Continuous variables presented as mean (SD); categorical variables as n (%). Missing data excluded; see Supplementary Table S3 for details.
Statistical significance: <sup>\*\*\*</sup> $p<0.001$ , <sup>\*\*</sup> $p<0.01$ , <sup>\*</sup> $p<0.05$ (2020 – 2022 vs 2016 – 2019); <sup>+++</sup> $p<0.001$ , <sup>++</sup> $p<0.01$ , <sup>+</sup> $p<0.05$ (2023 vs 2016 – 2019).
Abbreviations: EMS, emergency medical services; CVD, cardiovascular disease; COPD, chronic obstructive pulmonary disease; CKD, chronic kidney disease.

**Figure 1.** Study flow diagram of patients with ST-elevation myocardial infarction (STEMI) enrolled, 2016–2023. STEMI, ST-elevation myocardial infarction; EMS, emergency medical services.

### Unadjusted Temporal Trends in Pre-hospital Delay

Throughout the study period, women consistently experienced longer times from symptom onset to hospital arrival than men. The overall median pre-hospital delay was 170 minutes (IQR: 84–368) for women versus 124 minutes (IQR: 60–280) for men (*P*<0.001; see Supplementary Table S6 for percentile values). Encouragingly, pre-hospital delays decreased over time for both sexes. Figure 2 illustrates these trends: from 2016 to 2019, there were significant month-to-month declines in delay for both men and women (trend *P*<0.001), and after a plateau during 2020–2022, further improvements reappeared in 2023. No significant decline in delay was observed during the peak COVID-19 years (2020–2022), and a notable spike occurred in late 2022. Despite the overall shortening of delays in 2023 compared to 2016, the absolute sex gap in delay did not meaningfully narrow over time. In fact, the difference between female and male median delays showed a slight increasing trend from 2016 through 2019 (*P*<0.001 for trend), and it remained substantial in the subsequent years.

**Figure 2.** Trends in median pre-hospital delay (symptom onset to hospital arrival, minutes) by sex among all STEMI patients, 2016–2023. Vertical bars indicate median differences in delays between women and men. *P*-values represent tests for linear trends (*P* for trend) across the specified time periods.

Similar patterns were observed in the subset of patients arriving via EMS (Supplementary Figure S1). When examining components of delay separately, both the EMS call delay (symptom-to-call time) and the transportation delay (call-to-door time) showed overall temporal improvements parallel to the total pre-hospital delay (Supplementary Figures S2 and S3). Notably, however, no significant sex difference was found in EMS transport delay.

### Adjusted temporal trends of pre-hospital delay in women and men

After adjusting for age, cardiovascular risk factors, and comorbidities, women still had significantly longer pre-hospital delays than men across the entire distribution of delay times. Multivariable quantile regression confirmed that female sex was independently associated with longer pre-hospital delays across the entire distribution (Figure 3). For example, at the 50th percentile (median delay), women waited 24.44 minutes longer than men for hospital arrival (95% confidence interval [CI]: 22.71– 26.16; *P*<0.001). Importantly, this sex gap was even more pronounced at the upper tail of the delay distribution — by 2023, at the 90th percentile of delay, women’s times exceeded men’s by over 60 minutes. Similar findings were observed when comparing the 2020–2022 cohort to the 2016–2019 cohort.

**Figure 3.** Multivariable quantile regression of pre-hospital delay. The coefficient (95% CI) for women indicates the adjusted difference in delay between women and men during the reference period (2016–2019). The coefficient (95% CI) for period 2020–2022 or period 2023 reflects the change in delay among men relative to the reference period. The interaction term women × period represents the additional change in delay among women compared with men, relative to the reference period. Color coding: red indicates longer delays; blue indicates shorter delays. Significance levels: \*\*\**p*<0.001; \*\**p*<0.01; \**p*<0.05.

Overall pre-hospital delays improved significantly in the later periods for both sexes in adjusted analyses. Compared to the 2016–2019 baseline period, the adjusted median delay in 2023 was 13.99 minutes shorter (95% CI: 10.99–16.99) for men (indicating overall improvement), and in 2020–2022 it was 6.36 minutes shorter (95% CI: 3.80–8.91). An exception to this improvement was observed at the 90th percentile, where no significant decrease was found; in fact, delays were significantly longer in 2020–2022 compared to 2016–2019 (increase of 19.54 minutes, 95% CI: 2.46–36.61).

Crucially, the interaction term between female sex and admission period was statistically significant for all percentiles, indicating that the sex gap widened over time from 2016 to 2023, with the widening being most pronounced among patients with the most prolonged delays. These adjusted temporal trends were consistent in the EMS-transported population as well (no significant difference in patterns compared to the overall cohort).

### Subgroup Analyses

As shown in Figure 4, the female delay disadvantage was evident in every subgroup examined. Women had longer median pre-hospital delays than men across all age categories, hospital types, regions, and modes of transport. The magnitude of the sex gap varied somewhat, with the largest relative disparity observed in younger patients. In particular, women under 60 years old experienced the greatest delay difference compared to men of the same age group. Despite some differences in magnitude, the direction of the sex gap (women slower than men) did not change in any subgroup.

**Figure 4.** Subgroup analysis of median pre-hospital delay. The coefficient (95% CI) for women indicates the adjusted difference in delay between women and men during the reference period (2016–2019). The coefficient (95% CI) for period 2020–2022 or period 2023 reflects the change in delay among men relative to the reference period. The interaction term women × period represents the additional change in delay among women compared with men, relative to the reference period.

Temporal trends of delay improvement were generally similar across subgroups, with both women and men seeing shorter delays in 2023 than in 2016–2019. An exception was noted for patients arriving via EMS during the pandemic: in the EMS subgroup, pre-hospital delays in the 2020–2022 period were significantly longer than those in 2016–2019 for both sexes.

### Decomposition of Delay Components (EMS Call vs. Transport Times)

When we analyzed the two major components of pre-hospital delay separately (Figure 5), we found that the sex disparity was driven almost entirely by differences in the EMS call delay. Women waited significantly longer to contact EMS after symptom onset compared to men, whereas the difference in transport time (once EMS was activated) was minimal. For example, the adjusted median EMS call delay for women was about 13.42 minutes longer than for men (95% CI: 10.76–16.08; *P*<0.001). In contrast, the adjusted difference in EMS transportation time between women and men was only 1.49 minutes (95% CI: 0.81–2.17, *P*<0.001).

**Figure 5.** Multivariable quantile regression of EMS call delay and transportation delay. The coefficient (95% CI) for women indicates the adjusted difference in delay between women and men during the reference period (2016–2019). The coefficient (95% CI) for period 2020–2022 or period 2023 reflects the change in delay among men relative to the reference period. The interaction term women × period represents the additional change in delay among women compared with men, relative to the reference period. Color coding: red indicates longer delays; blue indicates shorter delays. Significance levels: \*\*\**p*<0.001; \*\**p*<0.01; \**p*<0.05.

## DISCUSSION

In this large national study of over 1.1 million STEMI patients in China, we identified several important findings. First, women consistently experienced significantly longer pre-hospital delays than men throughout the 2016–2023 study period, with a median delay difference of 46 minutes (even after accounting for clinical differences, the adjusted median delay was around 24 minutes longer in women). This female disadvantage was most pronounced in the youngest age group. Second, despite substantial overall improvements in pre-hospital times over the 8-year study period, the sex gap did not appreciably narrow. In fact, the difference between women’s and men’s delays remained large — and at the upper end of the delay distribution, the gap actually widened (by 2023, women in the 90th percentile of delay waited over an hour longer than men). Third, we found that this persistent disparity is primarily driven by delays in women calling EMS rather than differences in ambulance transport once help is activated. In other words, the critical “symptom onset to call” interval accounted for the vast majority of the sex delay gap, whereas EMS travel times were equivalent. Fourth, the COVID-19 pandemic was associated with a temporary reversal of progress: during 2020–2022, overall delays increased and the sex gap temporarily widened further, mainly due to prolonged decision times in women. These findings underscore that general gains in STEMI timeliness have not been shared equally by men and women.

Our finding of a sustained female delay disadvantage aligns with prior reports worldwide that women often take longer to seek medical help for STEMI symptoms, due to factors such as atypical symptom presentation, competing family/caregiver obligations, and lower perceived risk of heart attack. ^5,15,16^ However, the widening sex gap observed in our study contrasts with some recent data from other healthcare systems. For instance, a Swiss multi-center study reported that sex differences in pre-hospital delay narrowed after concerted system improvements and public campaigns, effectively reducing the gap between women and men.^11^ In China, despite broad improvements in processes of care through the Chest Pain Center accreditation, we did not see a similar closing of the gap. This suggests that China’s recent STEMI care improvements have benefited both sexes but may have been less effective for women, potentially because they did not directly address the specific barriers women face. The particularly large sex gap among patients with the longest delays (the 75th-90th percentiles) indicates that there remains a subgroup of women who experience severely prolonged times to treatment. These may be women facing compounded challenges (rural residence, low health literacy, etc.) who were not fully reached by general improvements, underscoring that systemic and social barriers disproportionately affect some women.

The overall reduction in median delays over the study period is encouraging and likely reflects the success of national quality efforts, notably the Chest Pain Center accreditation since 2016. The Chest Pain Center accreditation has standardized protocols, established regional coordination networks, and promoted public education on symptom recognition and timely EMS activation. Analysis of 1,372 Chest Pain Center — certified hospitals demonstrated that median door-to-balloon times decreased from 112 minutes in 2012 to 72 minutes by 2019, with 87% of cases in certified centers achieving the door-to-balloon ≤90-minute benchmark and top-performing centers consistently maintaining ≤50 minutes^17,18^. These systemic improvements plausibly contributed to the declining trend observed in our cohort, particularly during 2016–2019 and 2023.

By contrast, the stagnation and peak during 2020–2022 underscore the fragility of these gains during crises.^19^ The COVID-19 pandemic likely introduced new barriers and anxieties that disproportionately affected the patient decision-to-seek-care phase. Fear of COVID-19 exposure in healthcare settings, overwhelmed EMS systems, and lockdown-related communication barriers likely contributed to prolonged decision-making (call delay). EMS call delay is the primary driver of the sex gap (with minimal sex difference in transport delay). Women more frequently misattribute STEMI and consult family before calling EMS.^5^ Conversely, the neutral transport delay implies standardized EMS protocols effectively mitigate gender bias during transit — a success story of system-level optimization.

The amplified disparity among younger women contradicts traditional risk models. Potential explanations include: 1) under-40 women’s 3-fold higher angina misdiagnosis rates,^5^ 2) career-caregiving conflicts delaying help-seeking,^15,16^ and 3) younger men’s greater exposure to workplace first-aid training.^16^

The observed sex disparity in pre-hospital delay (median 46-minute difference) aligns with multinational registry data.^5,20^ The widening gap over time contrasts with some studies reporting stable differences, potentially reflecting unique pressures in our healthcare system or insufficiently targeted interventions for women.^11^ The critical role of call delay in driving the sex gap corroborates studies linking atypical presentation and care-seeking patterns to later EMS activation in women. Our observation of worsening call delay during the pandemic resonates with global reports of delayed STEMI presentations,^21^ but uniquely quantifies the differential impact on this specific delay component and on women.

Biological factors (atypical symptoms), psychological traits (altruistic hesitation), and structural barriers (childcare access) synergistically delay female care-seeking. Neuroimaging studies reveal gender differences in pain processing — women require higher ischemic thresholds to perceive symptoms as urgent.^16^ Institutionally, implicit bias persists; EMS dispatchers are 23% less likely to prioritize female chest pain calls.^5^ Pandemic stressors exacerbated these mechanisms through information overload and quarantine-induced social isolation.^22^

To address persistent gender disparities in STEMI care, targeted interventions must integrate biological and sociocultural dimensions. Public awareness campaigns should employ sex-specific symptom narratives (e.g., “sudden overwhelming fatigue” and “jaw numbness with nausea”) to challenge the misconception that heart disease primarily affects men, particularly targeting younger women aged <50 who exhibit higher mortality risks.^23^ These campaigns should leverage social media platforms frequented by women (e.g., TikTok, Xiaohongshu) and collaborate with community health workers to educate families on caregiving conflict resolution strategies.^9^

### Limitations

This study has several limitations that should be acknowledged. First, our analysis was based on a large hospital registry that is part of a voluntary quality improvement program. As a result, the participating hospitals (over 2,000 across China) may not be fully representative of all healthcare facilities in the country. In particular, tertiary-care centers and hospitals with resources to join the CPC program are likely overrepresented, while smaller rural hospitals or those not in the program may be underrepresented. This selection could limit the generalizability of our exact estimates of delay to all of China, although the core finding of a sex gap in delays is consistent with other settings. Second, the timing data for symptom onset and EMS call rely on patient self-report and clinical documentation, which introduces potential recall bias or recording error. Fourth, our registry did not capture the qualitative reasons for delay. We do not know, for instance, if a given patient delayed because they failed to recognize the severity of symptoms, because they attempted self-medication first, or because logistic issues (like lack of transportation or needing family assistance) interfered. Thus, our interpretations of “why” women delayed more are based on inference and external literature rather than direct survey data from these patients. Finally, the study period includes the COVID-19 pandemic, during which healthcare-seeking behavior was unusual; although we analyzed this period separately, the long-term trend may eventually resume its prior course, so the pandemic’s effects should be interpreted in context.

## CONCLUSIONS

In summary, this study demonstrates a persistent and concerning sex gap in pre-hospital delay for STEMI in China over an eight-year period. Women — especially younger women — continue to experience significantly longer times from symptom onset to hospital arrival than men, and this disparity, driven largely by delays in the decision-to-seek-care (EMS call), has not improved despite overall gains in system performance. These findings highlight that current improvements in STEMI care are not reaching men and women equally. Closing this sex gap through tailored public education, enhanced EMS training, and sex-specific quality metrics is essential.

## Nonstandard Abbreviations and Acronyms

CCA: Chinese Cardiovascular Association
EMS: Emergency medical service
NCPCP: National Cardiovascular Patient Care Program
MICE: Multiple Imputation by Chained Equations

## Acknowledgments

The authors thank all professionals in the Expert Committee and the Executive Committee of the China Chest Pain Center (https://www.chinacpc.org/home/auth/orgdesc) for their valuable contributions to this study.

## Supplemental Material

Table S1–S6

Figure S1–S3

## Data availability

The data, R code used in the analysis and original statistical outputs are available on reasonable request.

## Author contribution

LH and LS were responsible for funding acquisition. LH conceptualized the study. ZL, TZ, and YL contributed to the acquisition, analysis, and interpretation of data. ZL and TZ drafted the manuscript. HL, LH and LS provided critical revision of the manuscript. LH and LS provided administrative, technical, and material support. All authors were involved in reviewing the manuscript and approving the final version. All authors had access to all the data in the study and had final responsibility for the decision to submit for publication.

